# Evolution of Anti-SARS-CoV-2 IgG Antibody and IgG Avidity Post Pfizer and Moderna mRNA Vaccinations

**DOI:** 10.1101/2021.06.28.21259338

**Authors:** Kevin P. Bliden, Tiancheng Liu, Deepika Sreedhar, Jessica Kost, Jessica Hsiung, Su Zhao, Dingying Shan, Abira Usman, Naval Walia, Alastair Cho, Christophe Jerjian, Udaya S. Tantry, Paul A. Gurbel, Meijie Tang, Hongjie Dai

**Author notes:** These authors contributed equally (Kevin P. Bliden, Tiancheng Liu, Deepika Sreedhar, Jessica Kost).

## Abstract

Messenger RNA (mRNA) based vaccines (Pfizer/BioNTech and Moderna) are highly effective at providing immunity against severe acute respiratory syndrome coronavirus 2 (SARS-CoV-2). However, there is uncertainty about the duration of immunity, evolution of IgG antibody levels and IgG avidity (an index of antibody-antigen binding strength), and differences in the immune responses between vaccines. Here we performed a prospective pilot study of 71 previously COVID-19 free subjects upon receiving both doses of either the Pfizer (*n* = 54) or Moderna (*n* = 17) mRNA vaccine. Anti-spike protein receptor binding domain (RBD) IgG antibodies were measured longitudinally using a qualitative finger stick MidaSpot™ rapid test at the point-of-care for initial screening and a quantitative dry blood spot-based pGOLD™ laboratory test over ∼ four months post-vaccination. The average anti-RBD IgG antibody levels peaked at ∼ two weeks after the second dose of the vaccine and declined thereafter, while antibody avidity increased, suggesting antibody maturation. Moderna vaccine recipients compared to Pfizer vaccine recipients exhibited higher side effect severity, higher peak anti-RBD IgG antibody levels, and higher avidity up to the 90 days period. Differences in antibody levels diminished at ∼ 120 days post-vaccination, in line with the similar efficacy observed in the two vaccines. The MidaSpot™ rapid test detected 100% anti-SARS-CoV-2 RBD positivity for fully vaccinated subjects in both Pfizer and Moderna cohorts post full vaccination but turned negative greater than 90 days post-vaccination for 5.4% of subjects in the Pfizer cohort, whose quantitative anti-IgG were near the minimum levels of the group. Immune responses were found to vary greatly among vaccinees. Personalized longitudinal monitoring of antibodies could be necessary to assess the immunity duration of vaccinated individuals.

## Introduction

The severe acute respiratory syndrome coronavirus 2 (SARS-CoV-2) pandemic presents a great challenge to global health and has claimed millions of lives worldwide. Moreover, until herd immunity is successfully induced in a large population throughout the world, SARS-CoV-2 infections are likely to become endemic, leading to further loss of life. In response, multiple vaccine candidates based on RNA and DNA technologies, inactivated viruses, and other approaches have been rapidly developed^1-3^. Especially in the United States, two messenger RNA (mRNA) vaccines: BNT162b2 (Pfizer/BioNTech) and mRNA-1273 (Moderna), expressing SARS-CoV-2 spike glycoprotein, have received significant attention^4-6^. Early data analysis from the phase III Pfizer and Moderna vaccine clinical trials demonstrated that the vaccines were more than 90% effective in preventing illness and reducing disease severity and hospitalizations^7,8^. Further research has provided clear proof that both mRNA vaccines also elicited sufficient neutralizing activity against SARS-CoV-2 variants such as B.1.1.7, B.1.351, P.1, D614G, and their combinations^9,10^. Recent reports in Israel, Qatar, and the United States have further confirmed the effectiveness of mRNA vaccines against SARS-CoV-2 or its variants under real-world conditions^11-14^. Currently, tens of millions of doses are being administered daily worldwide under the FDA’s Emergency Use Authorization (EUA), with early longitudinal data suggesting that both mRNA vaccines are effective for at least six months and that a booster shot may be required^15^. There are many unknowns regarding the long-term efficacy of these newer vaccine technologies and the level and duration of protective immunogenicity. Furthermore, few studies have compared the currently available vaccines to assess potential differences in elicited antibody responses and side effects.

The production of antibodies in response to COVID-19 infection or vaccination in hosts is important to fight the SARS-CoV-2 virus. Various diagnostic and point-of-care (POC) platforms utilizing enzyme linked immunosorbent assay (ELISA)^16^, chemiluminescent immunoassays (CIA)^17^, and plasmonic gold near-infrared (NIR) fluorescence amplification^18^ have been developed to measure early IgA and IgM, and late IgG responses to the SARS-CoV-2 spike protein and its receptor-binding domain (RBD)^19,20^. Neutralization assays^21-25^ have been developed to quantify antibodies directly responsible for neutralizing viruses, and a strong correlation between neutralizing antibody titers with antibody levels against spike protein/RBD in COVID-19 patients has been reported^20,26-34^. In addition, antibody avidity measures the strength of antibody binding to viral antigens, shedding light on antibody maturation, and is potentially useful for assessing vaccine efficacy^35-40^. Therefore, it is important to measure both IgG levels and avidity to assess not only antibody quantity but also viral antigen binding ability post-infection or post-vaccination over time. On the other hand, reliable and inexpensive tests could be implemented in hospitals, clinics, various point-of-care (POC) sites, and even at home for frontline qualitative antibody detection to assess immune responses to COVID-19 infection or vaccination.

The primary objectives of this study were to longitudinally assess the immune responses to the Pfizer and Moderna mRNA vaccines by measuring anti-RBD IgG antibody levels and IgG avidity using the pGOLD™ assay. We also compared antibody production between the Pfizer and Moderna vaccines, and the relation to severity of post-vaccination reactions. We reported the results of longitudinal POC rapid testing of antibodies post-vaccination using the FDA EUA MidaSpot™ Antibody Combo Detection Kit^41^. Finally, for comparison we determined anti-RBD IgG and anti-NCP antibody levels in a cohort of healthy volunteers from the pre-pandemic era and from patients hospitalized with COVID-19.

## Results

### Demographic characteristics and safety

Between January and May 2021, a total of 71 adults vaccinated with Pfizer (*n* = 54) and Moderna (*n* = 17) were recruited for this study including Caucasians (63.4%), Asians (29.6%), and African Americans (7.0%), with a mean age of 46.8 ± 13.1 years. The common comorbidities reported were hypertension (16.9%), hyperlipidemia (8.5%), autoimmune type disorders (4.2%), and diabetes (1.4%) (Supplementary Table S1).

Post-vaccination local reaction and systemic events are included in Table 1. Myalgia, malaise, headaches, and fever were the most common symptoms, with increased frequency and severity after the second dose. After the first and second doses, respectively, 44% and 27% of subjects reported mild-to-moderate pain at the injection site. 60% of participants experienced mild-to-moderate systemic symptoms. Compared to participants who received the Pfizer vaccine, those who received the Moderna vaccine exhibited higher symptom frequency (78% vs. 55%) and severity for the 1^st^ and 2^nd^ vaccine dose (Figure 4c). One study subject receiving the Moderna vaccine reported syncope and vertigo, requiring a short hospitalization. Symptoms generally peaked at two days after vaccination and resolved within a week. No subjects experienced recurring symptoms or contracted COVID-19 after the second dose.

**Table 1.** Vaccine side effects and severity. The table presents the percentage of participants who reported the listed side effects. Participants could have listed one or more of the possible side effects. The table also presents the average (+/-standard deviation) self-reported side effect severity on a scale of 0 (low) to 10 (high). The most common reported side effect after two doses was soreness of arm, with Moderna subjects reporting a higher percentage than Pfizer subjects. A higher percentage of Pfizer subjects reported no side effects after two doses than Moderna subjects. Only one participant was hospitalized, after the first Moderna vaccine dose. Overall, side effect severity ratings were higher for Moderna subjects than Pfizer subjects.

|  | Total Vaccinees |  | Pfizer |  | Moderna |  |
| --- | --- | --- | --- | --- | --- | --- |
|  | (n=71) |  | (n=54) |  | (n=17) |  |
|  | 1st Dose | 2nd Dose | 1st Dose | 2nd Dose | 1st Dose | 2nd Dose |
| Side Effects (%) |  |  |  |  |  |  |
| Arm Soreness | 52.1% | 28.2% | 46.3% | 22.2% | 70.6% | 47.1% |
| Myalgia | 2.8% | 1.4% | 1.9% | 1.9% | 5.9% | 0.0% |
| Fatigue/Malaise | 21.1% | 36.6% | 16.7% | 33.3% | 35.3% | 47.1% |
| Body Aches/Joint Pain | 5.6% | 16.9% | 3.7% | 13.0% | 11.8% | 29.4% |
| Nausea/Diarrhea | 8.5% | 2.8% | 9.3% | 1.9% | 5.9% | 5.9% |
| Headache | 12.7% | 22.5% | 11.1% | 20.4% | 17.6% | 29.4% |
| Fever | 7.0% | 26.8% | 5.6% | 20.4% | 11.8% | 47.1% |
| Chills | 5.6% | 19.7% | 5.6% | 16.7% | 5.9% | 29.4% |
| Cough/Dry Mouth | 1.4% | 0.0% | 1.9% | 0.0% | 0.0% | 0.0% |
| Dizziness/Syncope | 2.8% | 1.4% | 1.9% | 1.9% | 5.9% | 0.0% |
| Flu-Like Symptoms | 0.0% | 2.8% | 0.0% | 1.9% | 0.0% | 5.9% |
| Hospitalization | 1.4% | 0.0% | 0.0% | 0.0% | 5.9% | 0.0% |
| None | 25.4% | 18.3% | 31.5% | 24.1% | 5.9% | 0.0% |
| **Severity Scale (0-10)** | **2.7 +/- 2.6** | **3.6 +/- 2.6** | **2.3 +/- 2.5** | **3.1 +/- 2.6** | **3.9 +/- 2.8** | **5.2 +/- 2.1** |

### Evolution of anti-RBD and anti-NCP IgG levels post-vaccination

To investigate the immune response to SARS-CoV-2 mRNA vaccines, we measured pre-first dose (day 0) and vaccine-elicited anti-RBD IgG antibodies at various times post-vaccination in 71 vaccinees between 19 and 146 days following the first vaccine dose. Based on the days from first-dose injection, vaccinated samples were divided into five groups: D0 (0 days, prior to first dose), D19-28 (20 days for Pfizer vaccine; 27 days for Moderna vaccine; prior to second dose), D33-55 (33-55 days post-first dose), D61-83 (61-83 days post-first dose), D89-108 (89-108 days post-first dose), and D112-146 (112-146 days post-first dose).

Rapid testing was performed as a preliminary qualitative detection of antibodies elicited by vaccination with a 10 μL fingerstick sample, meanwhile a 10 μL DBS sample was saved for quantitative pGOLD™ assay. The POC MidaSpot™ lateral flow assay detected anti-RBD IgG in ∼ 90% of vaccinees after the first dose (Table 2, Figure 1) and 100% positive rate in fully vaccinated individuals at day 33-55 for both the Moderna and Pfizer cohorts. With ∼ 10% vaccinees, MidaSpot detected low or negative IgG signals after the first dose but detected clear IgG lines after the second dose (Figure 1a, 1c). The negative agreement is 100% for both vaccines in the pre-vaccination group (Table 2).

**Table 2.** Serial testing of vaccinated individuals on the Nirmidas MidaSpot™ Rapid Antibody Test. The tables comprise a total of 71 individuals, of whom 17 received the Moderna vaccine and 54 received the Pfizer vaccine. Of those who received the Pfizer vaccine, 75.9% of individuals (41/54) returned for serial testing and had two or more data points in this table. Of those who received the Moderna vaccine, 64.7% of individuals (11/17) returned for serial testing and had two or more data points in this table. The 21 or 28 days from first vaccine dose was collected from the participants before the second dose of the vaccine. All collections after the 21- or 28-day timepoint were done after the subject received the second dose of the vaccine.

| Moderna |  |  |  |
| --- | --- | --- | --- |
| Days from Vaccination | Total Samples | IgG + | IgG NPA |
| -1 - 0 | 5 | 0 | 100.0% |
| Days from Vaccination | Total Samples | IgG + | IgG PPA |
| 19 - 28 | 9 | 8 | 88.9% |
| 33 - 55 | 8 | 8 | 100.0% |
| 61 - 83 | 10 | 10 | 100.0% |
| 89 - 108 | 6 | 6 | 100.0% |
| 112 - 144 | 6 | 6 | 100.0% |
| Overall | 39 | 38 | 97.4% |

| Pfizer |  |  |  |
| --- | --- | --- | --- |
| Days from Vaccination | Total Samples | IgG + | IgG NPA |
| -1 - 4 | 19 | 0 | 100.0% |
| Days from Vaccination | Total Samples | IgG + | IgG PPA |
| 19 - 28 | 23 | 18 | 78.3% |
| 33 - 55 | 27 | 27 | 100.0% |
| 61 - 83 | 40 | 39 | 97.5% |
| 89 - 108 | 9 | 9 | 100.0% |
| 112 - 144 | 37 | 35 | 94.6% |
| Overall | 136 | 128 | 94.1% |

**Table 2. Serial testing of vaccinated individuals on the Nirmidas MidaSpot™ Rapid Antibody Test.**
| Combined |  |  |  |
| --- | --- | --- | --- |
| Days from Vaccination | Total Samples | IgG + | IgG NPA |
| -1 - 4 | 24 | 0 | 100.0% |
| Days from Vaccination | Total Samples | IgG + | IgG PPA |
| 19 - 28 | 32 | 26 | 81.3% |
| 33 - 55 | 35 | 35 | 100.0% |
| 61 - 83 | 50 | 49 | 98.0% |
| 89 - 108 | 15 | 15 | 100.0% |
| 112 - 144 | 43 | 41 | 95.3% |
| Overall | 175 | 166 | 94.9% |

**Figure 1.**
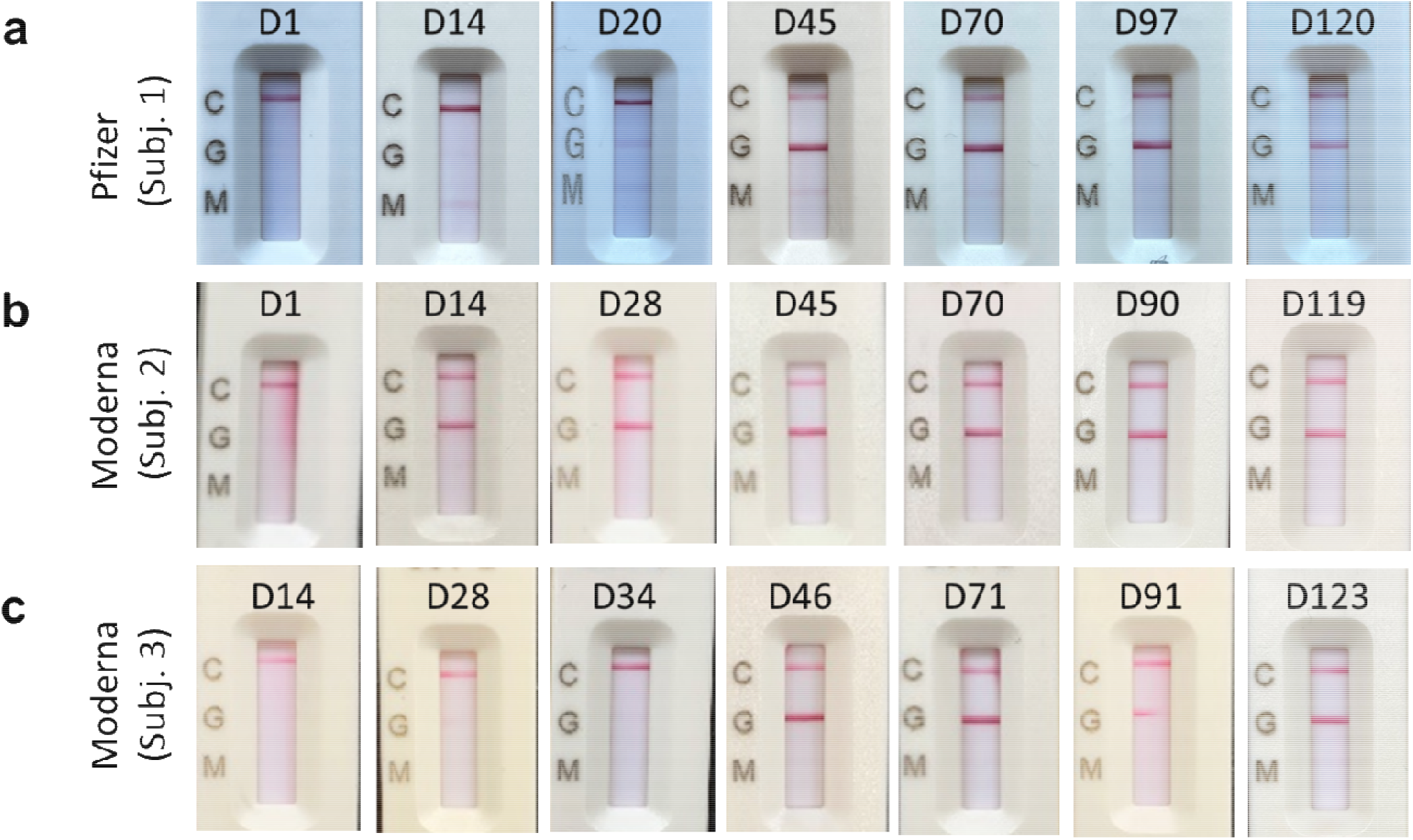
Representative longitudinal MidaSpot ™ rapid antibody test results fo vaccinated subjects. (**a**) MidaSpot ™ results from Day 1 to Day 120 for a Pfizer vaccinee subject 1. IgG signal (G) was visible/positive on Day 20 post the first dose and became high positive after the second vaccine dose tested from D45 to D120. (**b**) MidaSpot results from Day 1 to Day 90 for a Moderna vaccinee subject 2. IgG signal increased turned to high positive on Day 14 post the first dose and remained high positive over time. (**c**) MidaSpot results from Day 14 to Day 123 for a Moderna vaccinee subject 3. IgG signal was visible on Day 28, then disappeared on Day 34, and increased obviously after the second dose and remained high positive. Day 91 data showed an interrupted signal line likely due to a defective test card, but the line was clearly visible.

Using the pGOLD™ assay, we detected anti-RBD IgG with a 100% positive rate for all vaccinees at all the post-first dose timepoints (D19-28, D33-55, D61-83, D89-108, and D112-146), and a 100% negative rate in the pre-vaccination group (D0), as well as pre-pandemic samples. Since the mRNA vaccines encode the SARS-CoV-2 spike protein, the pGOLD™ assay only detected positive anti-RBD for the vaccinees without detecting anti-NCP IgG (Figure 2a, 2c, 2d), compared to both positive anti-RBD and anti-NCP detected in COVID-19 patients (Figure 2b, 2e, 2f). It was also observed that there was a clear increase in baseline signals for anti-NCP IgG after the D33-55 timepoint, but still below the cutoff value of 10.3 (Figure 2d). However, the reason is unknown.

**Figure 2.**
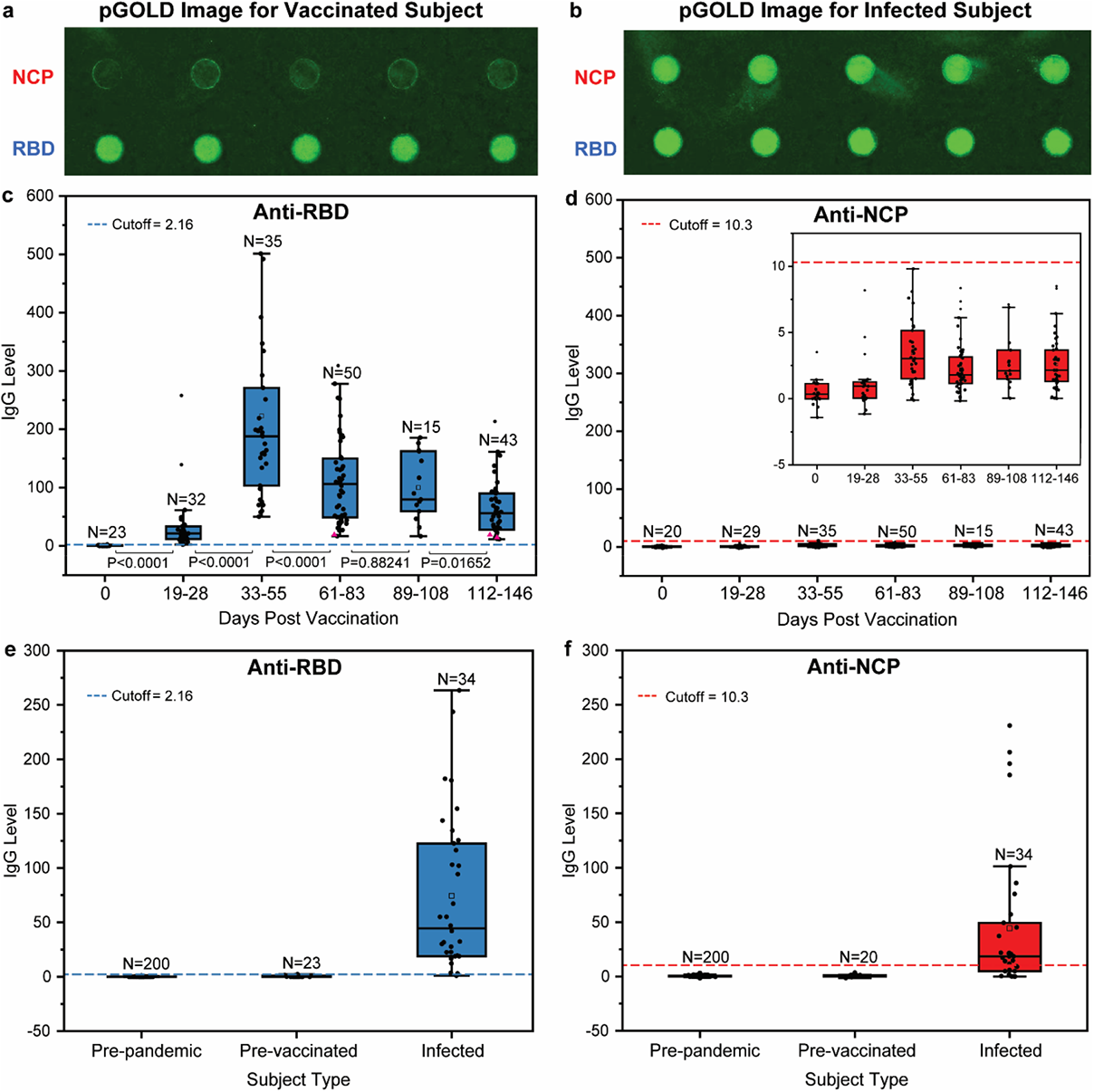
Quantitative pGOLD™ anti-RBD, NCP IgG Levels in vaccinated and infected subjects. (**a**) Confocal fluorescence scanned images of IgG levels against RBD and NCP antigens (printed circular spots on pGOLD™) acquired after testing a DBS sample of a vaccinated subject. Low anti-NCP baseline signal was observed. (**b**) Same as (a), but with a sample from a SARS-CoV-2 infected patient showing both high anti-NCP IgG and anti-RBD IgG signals. (**c**) Box plots for anti-RBD IgG levels for vaccinated subjects in a range from Day 0 (pre-vaccination), Day 19-28 (before the second vaccine dose), Day 33-55, Day 61-83, Day 89-108, and Day 112-146, *n* = 198. Cutoff = 2.16. Two samples are outside the displayed range. Samples that are negative on MidaSpot™ are shown as pink triangles at the minimum of antibody level. (**d**) Same as (a), but for anti-NCP IgG levels. *n* = 192. Cutoff = 10.3. Inset: a zoom in view of the low anti-NCP IgG levels all below cutoff but showing a slight variation post vaccination. (**e**) Box plots showing anti-RBD IgG levels of SARS-CoV-2 of pre-pandemic subjects (serum samples, *n* = 200) below cutoff, those of pre-vaccinated subjects (Day 0, DBS samples, *n* = 23) below cutoff. In contrast, most of infected subjects (DBS samples, *n* = 34) above cutoff. (**f**) Same as (c) but for anti-NCP IgG levels. SARS-CoV-2 pre-pandemic subjects (serum samples, *n* = 200) showing anti-NCP below cutoff, similar pattern to pre-vaccinated subjects (Day 0, DBS samples, *n* = 20). Infected subjects (DBS samples, *n* = 34) are most above cutoff.

The quantitative pGOLD™ assay data showed an anti-RBD IgG peaking and then declining trend in all vaccinated subjects (Figure 2c). The anti-RBD antibodies evolved from a low baseline level (D0), upward to mildly positive (D19-28), rapidly peaking (D33-55), and then gradually decreasing (D61-83, D89-108, D112-146) (Figure 2c). Each group’s general data information is summarized in Supplementary Table S2. The normality test by the Shapiro-Wilk method showed that the anti-RBD IgG levels grouped by time points were non-normal for D0, D19-28, D33-55, D61-83, and D112-146, and normal for D89-108 (Supplemental Table S3). It is to be noted that the D89-108 timepoint only had 15 samples. We then used a nonparametric statistical analysis method, the Mann-Whitney test (Supplementary Table S4), to analyze the similarity between anti-RBD IgG levels measured at various timepoints. The anti-RBD IgG levels in the pre-vaccination (D0) and pre-pandemic groups (200 serum samples, collected before November 2019) were much lower than the post-vaccination groups (D19-28, D33-55, D61-83, D89-108, D112-146, and COVID-19 positive) with *p* values in the range of 0.0001 and 0 (Supplementary Table S4). The anti-RBD IgG levels measured at 19-28 days (D19-28 group), before the second dose, were significantly lower than the later groups (D33-55, D61-83, D89-108, and D112-146) with *p* value < 0.0001, respectively (Supplementary Table S4). The anti-RBD IgG measured at 33-55 days (D33-55 group) exhibited the highest levels with *p* value < 0.0001 (D19-28 vs. D33-55) and *p* value < 0.0001 (D33-55 vs. D61-83). The anti-RBD IgG levels measured at two, three, and four months (D61-83, D89-108, and D112-146) were close, with *p* values: 0.88241 (D61-83 vs. D89-108) and 0.01652 (D89-108 vs. D112-146), suggesting a small decrease after two months (Supplementary Table S4). Our results also confirmed that mRNA vaccine-elicited median anti-RBD IgG levels measured after the second dose were higher than those in COVID-19 with *p* value < 0.0001 (D33-55 vs. COVID-19) (Figure 2c vs. 2e, Supplementary Table S4).

### Evolution of anti-RBD IgG avidity post vaccination

A SARS-CoV-2 anti-RBD IgG avidity assay was developed on Nirmidas’ pGOLD™ platform to investigate antibody-antigen binding strength and stability under denaturing conditions. In the pGOLD™ anti-RBD IgG avidity test, a 4 M urea-containing sample buffer and urea-free normal sample buffer were employed to dilute the same sample solution derived from a vaccinee’s DBS, then loaded into neighboring wells on pGOLD™ biochips for detecting anti-RBD IgG with and without urea treatment (Figure 3a). A higher avidity index (the ratio of IgG level with urea treatment over that without urea treatment) indicated that the detected antibody possessed a stronger and more stable binding to the antigens on pGOLD™ against denaturing forces. Averaged over all participants in the two mRNA vaccinated cohorts, we observed that the anti-RBD IgG avidity showed a generally increasing trend with time (Figure 3a), indicating maturation of antibodies elicited by mRNA vaccines. We also analyzed anti-RBD IgG antibodies measured under 4 M urea denaturing conditions and observed smaller decreases over time (Figure 3b) than the overall anti-RBD antibody levels measured without urea treatment (Figure 2c), revealing a more steadily increasing level of strong RBD binding antibodies over time (Figure 3b).

**Figure 3.**
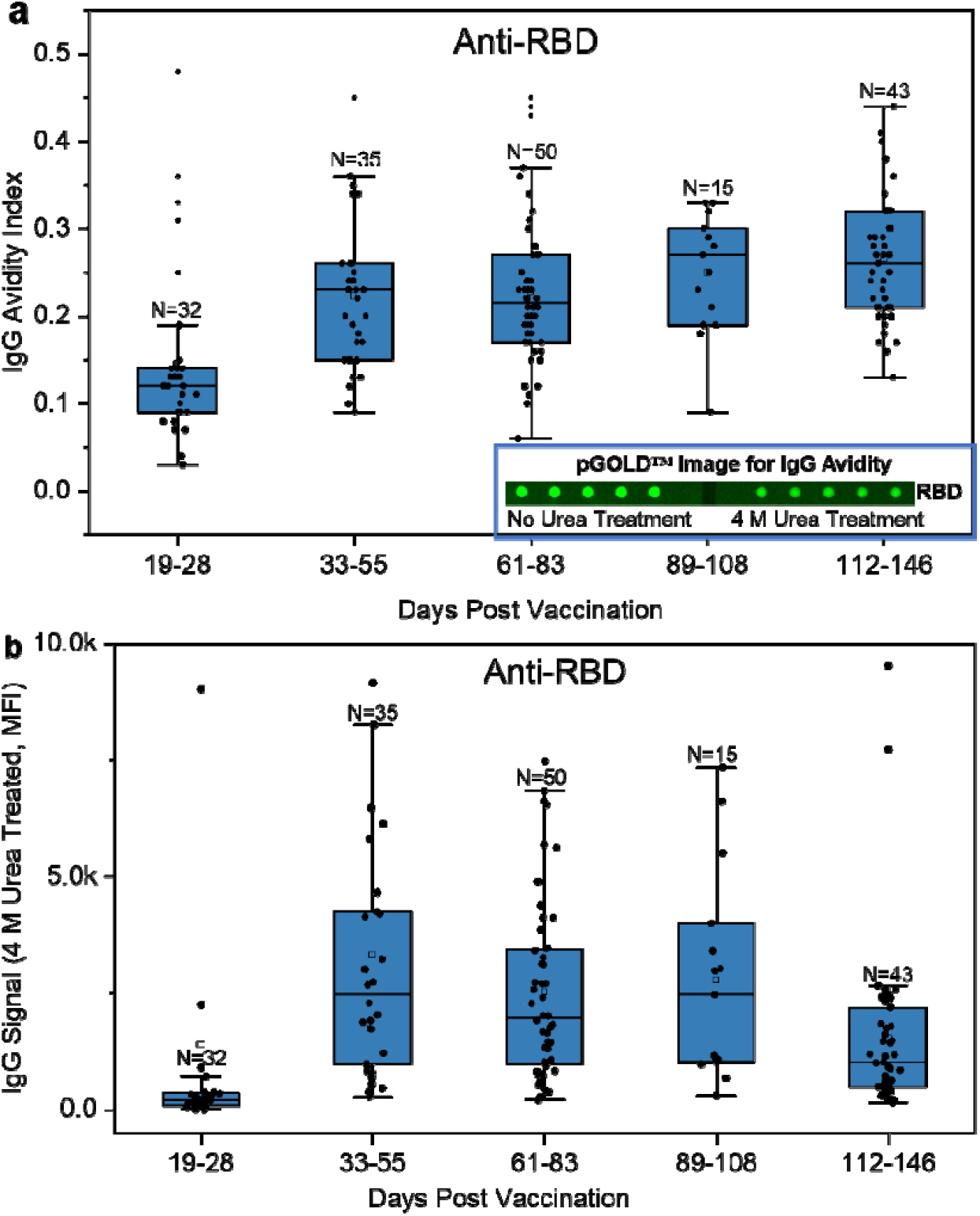
The kinetics of anti-RBD IgG avidity over time post vaccination. (**a**) Anti-RBD IgG avidity index for all vaccinated subjects showed an increasing trend over time (unit for x-axis: days post the first dose). *n* = 175. Inset: pGOLD™ fluorescence images showing anti-RBD signals on 5 antigen spots for the same vaccinee DBS sample obtained with (right) and without (left) urea treatment. The fluorescence intensity is weaker in the urea treatment case, and avidity index is measured as the ratio of the signal (in MFI, Median Fluorescence Intensity) of the urea-treated case to no-treatment case. (**b**) Box plots of fluorescence Intensities (MFI) of anti-RBD levels measured with 4 M urea treated DBS samples from vaccinees over time. The plots revealed there were small changes at timepoints (D33-55, D61-83 and D89-108).

**Figure 4.**
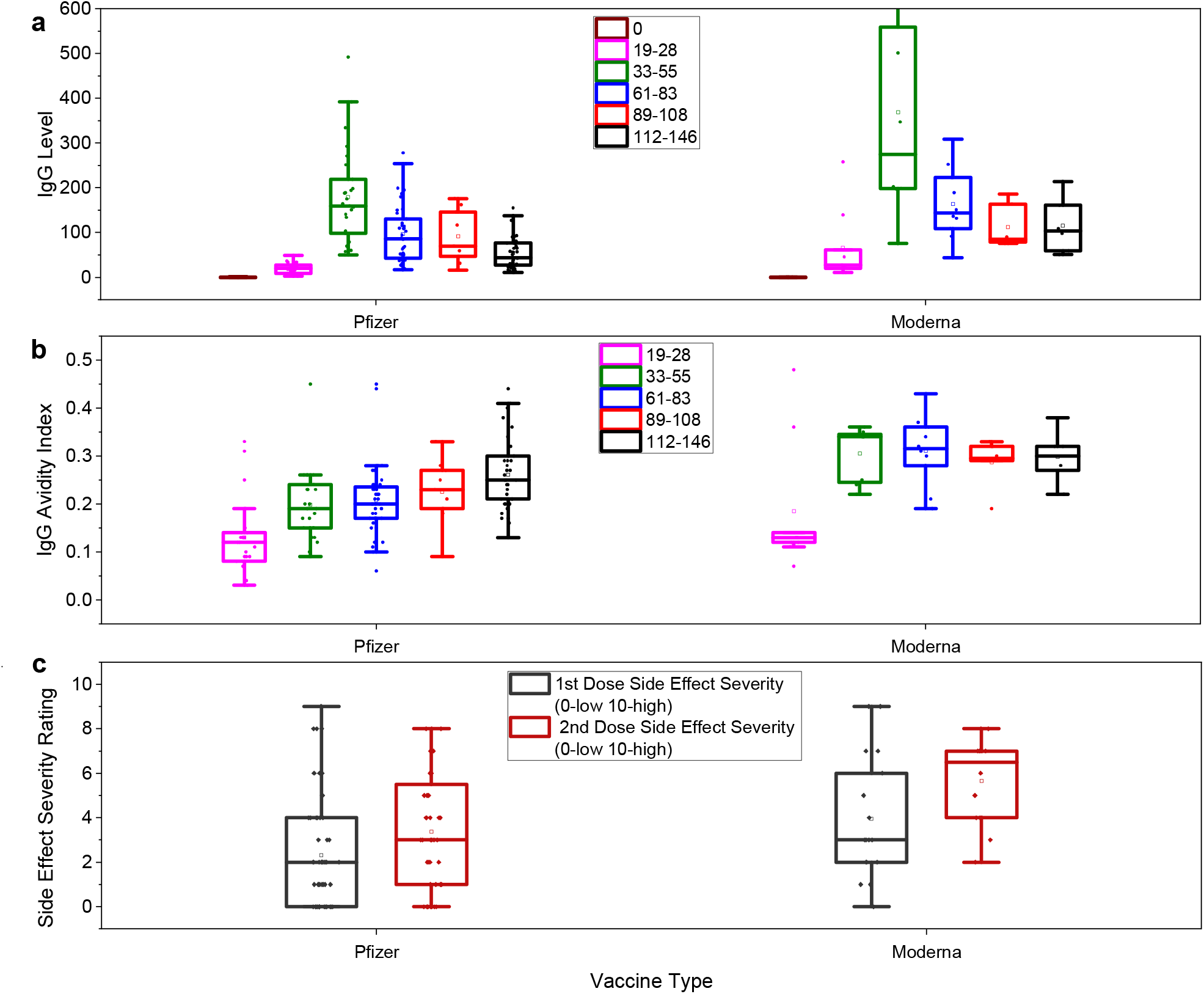
Different responses between Pfizer- and Moderna-vaccinated cohorts. (**a**) anti-RBD IgG levels measured in the Pfizer (54/71=76.1%) and Moderna vaccinees (17/71=23.9%), trended to increase from baseline (D0) until Day 33-55 and decreased henceforth for both vaccinated cohorts. The Moderna subjects had a higher peak than Pfizer subjects at the Day 61-83 timepoint, but the last timepoint (D112-146) indicated closer IgG levels for both. The text box in the middle inset of the graph indicates days post the first vaccine dose. (**b**) Box plots of anti-RBD IgG avidity index for the Pfizer vaccinee subjects showed an increasing trend, and Moderna subjects showing a leveling trend, with similar final avidity indices for vaccinated cohorts at timepoint D112-146. (**c**) Self-reported side effect severity ratings showed higher median severity for Moderna subjects, after both doses.

### Immune responses of Pfizer versus Moderna vaccinated cohorts

We then analyzed the immune responses of the Pfizer and Moderna vaccinated cohorts separately. We observed that the Moderna vaccinees showed higher average anti-RBD IgG levels in the first three months than the Pfizer vaccinees (Figure 4a), accompanied by more intense side effects right after the first and second dosing (Table 1, Figure 4c). The differences in the average anti-RBD IgG levels measured with the two cohorts decreased over time and were on par measured at the ∼ four-month timepoint (Figure 4a).

The average anti-RBD IgG avidity index measured for the Moderna cohort showed a jump after the second dose (a similar trend as the antibody level) to a level much higher than the Pfizer cohort in the same time course (Figure 4b). While the Moderna cohort’s mean anti-IgG avidity index remained relatively stable (Figure 4b) over four months, the Pfizer cohort of vaccinees showed a clear increasing trend in avidity, reaching a similar average as the Moderna cohort but still with a lower minimum avidity (Figure 4b).

We also observed that 100% of the Moderna cohort of vaccinees tested positive by the qualitative MidaSpot™ rapid test after receiving the first dose through the four-month period of monitoring (Table 2). On the other hand, 5.4% of the Pfizer vaccinees turned negative by rapid test greater than three months after the first dose. Close comparison with quantitative pGOLD™ antibody data showed that the anti-RBD levels in these ‘negative’ samples were still above the cutoff value relative to the pre-vaccination level but were indeed close to the minimum levels of the corresponding groups of the cohort, and at least three-fold lower than the median levels (Figure 2c, Supplementary Table S5).

## Discussion

The SARS-CoV-2 spike protein is a homo-trimeric glycoprotein containing the receptor binding domain (RBD), which mediates virus infection to host cells by interacting with its receptor: human angiotensin converting enzyme 2 (ACE-2) on the cell surface^19^. Blocking the interaction between RBD and ACE-2 can efficiently prevent SARS-CoV-2 infection. Most neutralizing antibodies in convalescent COVID-19 patients have been identified to target spike protein RBD epitopes at or near the ACE2/RBD binding interface^20,25,26^. Both the Pfizer BNT162b2 and Moderna mRNA-1273 vaccines were designed to express the full-length SARS-CoV-2 spike protein in a prefusion state to induce a sustained humoral immune response in vaccinated individuals^3,42,43^. The mRNA vaccines have shown high efficacy in the real world and are largely responsible for bringing COVID-19 under control in the US^13,14^ and other regions^11,12^. It is currently unknown how long immunity will last, and booster vaccine shots in about one year from the initial dose have been suggested/forecasted.

Immunity to COVID-19 and other infectious diseases are dependent on the generation of systemic neutralizing antibodies in the host elicited by past infection or vaccination. Strictly speaking, neutralizing antibody levels in vaccinees should be closely monitored longitudinally for a large cohort of vaccinees to assess the duration of immunity to COVID-19. However, standard neutralization assays^21,22^ require live viruses, special facilities, and are costly and time-consuming, making them difficult to implement in a large vaccinated population. Importantly, there has been accumulating evidence that anti-spike/RBD IgG levels are strongly correlated with neutralizing activity in COVID-19 patients^26-30^ and vaccinated individuals^44^. It is reasonable that anti-RBD IgG levels are measured to assess neutralizing antibody levels, which could be done quantitatively in many laboratories or qualitatively at POC sites. In this study, we focused on measuring anti-RBD IgG levels and anti-RBD IgG avidity to investigate the kinetics and maturation of vaccine-elicited antibodies.

One of our findings was that both the Pfizer and Moderna mRNA vaccines elicited anti-RBD IgG effectively in 100% of the 71 vaccinees studied longitudinally. The average IgG levels were boosted by the second dose and peaked after ∼ two weeks. Antibody levels then showed a downward trend up to the four-month timepoint. These observations were similar to recent reports on the Pfizer and Moderna vaccines^5,15,45^. Comparing these two cohorts side-by-side revealed that the average antibody levels in the Moderna cohort in the first three months post-vaccination were higher than in the Pfizer cohort, consistent with the more severe, but mostly tolerated side effects (with only one hospitalization in the Moderna cohort). However, at the four-month timepoint, anti-RBD IgG levels in the Moderna and Pfizer cohorts were closer, suggesting potentially similar long-term effects and vaccine efficacy.

The antibody avidity index provides an assessment of antibody-antigen binding strength by measuring antibodies specifically bound to antigens in the presence of a denaturing agent. It is known that the antibodies produced in the early days of an infection show low avidity with weak binding to the viral antigen. Avidity increases as antibodies mature over months or years through processes including colonial expansion, hypermutation, and affinity selection in the germinal center^46^. Antibody avidity has been used to assess recent vs. remote infections^18,35-39^ and to analyze vaccination efficacy, immunity, and the success of convalescent plasma-based antibody therapy^47-49^.

Overall, the average avidity index of the two mRNA vaccines showed an increasing trend over the four months period monitored (Figure 3b). Consistent with a stronger immune response, the average anti-RBD IgG avidity for the Moderna cohort was higher than that of the Pfizer cohort but did not show an obvious upward trend in the 4-month period. In contrast, anti-RBD IgG in the Pfizer cohort showed a steady increase approaching that of the Moderna cohort, suggesting a clear antibody maturation process. In the four-month post-vaccination timepoint (Figure 4a, 4b), the two mRNA vaccines were associated with similar antibody levels and avidity (slightly higher for Moderna), consistent with the similar ∼ 95-98% efficacy (also slightly higher for Moderna) in protection from COVID-19 infection from clinical trials.

Both antibody concentration/quantity and antigen binding strength/avidity are important to immunity and can be used to analyze vaccine effectiveness. As antibody concentration wanes over time post-vaccination, a higher avidity could be important to sustain immunity and maintain the ability to fight viral infection at reduced antibody levels. We clearly observed increasing anti-RBD avidity and maturation with the Pfizer vaccinee cohort (Figure 4b), a welcoming result since even though the antibody concentration decreased over time, the viral-antigen binding strength increased for the remaining antibodies. The Moderna vaccinee cohort showed high antibody levels and avidity in the first 4 months period with more severe side-effects, without an obvious increase in avidity. The differences between the two mRNA vaccines were interesting but not understood currently. Clearly, it is important to continue monitoring the two cohorts over longer periods of time to investigate further evolutions of antibody levels and avidity and their relations to immunity. It is possible that antibody maturation continues to further boost avidity and afford useful viral fighting capabilities even at low antibody levels.

We observed that women and subjects < 50 years of age tended to exhibit a better immune response, and subjects with a history of autoimmunity had a lower immune response. By one-way ANOVA analysis, we analyzed subjects that were ≤ 50 years and > 50 years of age. Anti-RBD IgG was significantly higher at the three-to four-months measurement point (75.5±49.2 vs. 48.5±34.0, p=0.047). We analyzed subjects who had autoimmunity (*n* = 4) and found that the anti-RBD IgG levels were significantly lower at the three-to four-months point compared to the rest of the group (*n* = 40) (75.5±49.2 vs 48.5±34.0, p=0.045).

We performed the MidaSpot™ POC rapid antibody test as an initial qualitative assessment of immune response to vaccination, during which dried blood spots were collected for quantitative pGOLD™ assay. The results also shed light on whether rapid antibody tests could be used to produce meaningful frontline assessment of antibody levels elicited by the vaccines at POC sites. Importantly, the FDA EUA MidaSpot™ rapid antibody test detected anti-RBD IgG with 100% sensitivity in fully mRNA vaccinated subjects (∼ two weeks after the second dose). For the Moderna vaccinees, 100% antibody positivity persisted to the ∼ four-month timepoint, with ∼ 94% positivity for the Pfizer cohort. Importantly, the 5.4% negative results were from the three-to four-month timepoints, for vaccinees whose quantitative anti-RBD levels measured by pGOLD™ were close to minimum levels of the corresponding groups of the cohort, and at least three-fold lower than the median values (Table 2, Supplementary Table S5). We conclude that the MidaSpot™ rapid test can indeed provide a useful qualitative, preliminary assessment of immune response to vaccination. For subjects whose MidaSpot™ rapid test anti-RBD result turned negative or very faint in the signal line, the corresponding quantitative antibody level was low and warranted further testing or clinical decisions.

The pGOLD™ assay is a novel nanotechnology assay platform capable of quantifying antibody levels and binding affinity to viruses^18,39,50-52^. It is also capable of detecting antibodies in DBS or saliva samples and could offer a non-invasive approach to assess antibody response to vaccination. Furthermore, the combination of the MidaSpot™ rapid antibody test and the pGOLD™ assay could generate more complete information on vaccine-elicited antibody levels by quantity and quality with time.

Limitations of our current study include the small sample size that is unequal among groups, missing data at some timepoints, and the limited population diversity.

## Conclusions

In summary, we performed a longitudinal study of two prominent mRNA vaccines for SARS-CoV-2 by rapid antibody testing and quantitative measurements of antibody levels and binding strength based on the avidity index. The 4-months post-vaccination results are presented here as an interim report, and the study will continue up to more than a year. Thus far, both vaccines elicit similar levels of anti-RBD IgG and avidity at the 4-month timepoint, suggesting that both vaccines are similarly efficient at protecting vaccinated populations. We compared the two mRNA vaccinee groups and observed interesting differences, and the results might shed light on vaccine development. An ideal vaccine would be one that generates not only high antibody levels, but also high viral-antigen binding strength, and at the same time affords well-tolerated side effect severity. With the vaccinees, we observed that in general, although antibody levels decreased over time, antibody maturation was observed with increased antigen binding strength, boding well for sustained immunity. We did observe highly variable immune responses including those with well below average anti-RBD IgG levels and avidity. It is therefore important to monitor immune responses at the individualized and personalized level, identify those who are still at risk even after vaccination, and provide meaningful measures to protect them from infections.

## Data Availability

Data are available in main manuscript and Supplementary information.
Further requests for materials should be addressed to P.A.G, M.T. and H.D.

## Methods and Materials

### Prospective study design

The study (NCT# 04910971) was performed in accordance with standard ethical principles and approved by the LifeBridge Health local Institutional Review Board under Protocol # 1707882. All participants provided written consent. Subjects ≥ 18 years of age receiving SARS-CoV-2 vaccination with no recent or previous COVID-19 infection history, or those detected negative by MidaSpot™ COVID-19 antibody rapid test, were enrolled in the study. For each participant, two 10 µL blood samples were collected by fingerstick and applied to a dry blood spot (DBS) card and a MidaSpot™ lateral flow test card respectively before the administration of vaccines (0 day), and at 19-28 (pre-second dose of vaccination), 33-55, 61-83, 89-108, and 112-146 days after the initial vaccine dose. Demographics and medical history were collected at screening, and vaccination side effects (type and severity) were collected and recorded using a questionnaire. For comparative analysis, anti-RBD IgG and anti-NCP antibody data in a cohort of healthy volunteers from the pre-pandemic era (*n* = 200), and COVID-19 patients hospitalized (*n* = 34) from a previous study (NCT# 04493307) confirmed by positive reverse transcription polymerase chain reaction (RT-PCR) tests were included.

### Antibody testing methods

Nirmidas’ MidaSpot™ COVID-19 Rapid Antibody Combo Detection Kit was used for initial screening and assessment of immune response (Figure 1). This test was approved by the United States Food and Drug Administration (FDA) Emergency Use Authorization (EUA) for POC testing, ideally suited for qualitative antibody testing at any healthcare site. The lateral flow assay detected both immunoglobulins IgG and IgM against the SARS-CoV-2 viral spike protein receptor-binding domain (RBD), which is the same antigen expressed by the mRNA vaccines. In addition, a dry blood spot card for each subject taken at each timepoint was stored at -20 ^°^C, batched, and shipped to Nirmidas Biotech Inc. (Palo Alto, CA, USA) to perform the pGOLD™ COVID-19 High Accuracy IgG/IgM Assay to quantify antibody levels and avidity as previously described, with minor modifications^18^. Briefly, we conjugated iFluor™820 succinimidyl ester (ATT Bioquest, Cat. No:1399) and CF647 succinimidyl ester (Millipore Sigma, SCJ4600048) to anti-human IgG and anti-human IgM, respectively. The iFluor™820-labeled anti-human IgG and CF647-labeled anti-human IgM were used for two-color simultaneous multiplexed detection of IgG and IgM against RBD and nucleocapsid protein (NCP) antigens printed on pGOLD™ slides using a robotic micro-arrayer.

### Processing and preparation of DBS samples for the pGOLD™ assay

A 2 mm circle was punched out from each DBS card (comprised of 10 μL of fingerstick whole blood drop, dried on the card) and dissolved into 160 µL sample solution containing 10% fetal bovine serum (FBS), 0.05% Tween 20 and 0.1% NaN_3_ in 1X phosphate-buffered saline (PBS) under 1 hour shaking at room temperature. After a brief centrifugation, the DBS-dissolved sample solution was heated for 30 minutes at 56 ^°^C. The heat-inactivated solution was used for the pGOLD™ assay immediately or stored at 4 ^°^C for later use.

### Multiplexed pGOLD™ antibody assay against spike protein RBD and nucleocapsid protein

The pGOLD™ antibody assays were performed in a 64-well format in the following steps. **1)** Blocking: All wells were blocked with a blocking buffer containing 2% bovine serum albumin (BSA) in 1X PBS for 30 minutes at room temperature; **2)** Sample incubation: each well was then incubated with 100 µL of 2X diluted dissolved DBS sample solution in 10% FBST (10% FBS including 0.05% Tween 20) for 60 minutes at room temperature. A mixture of diluted anti-RBD IgG/IgM purified antibodies as signal normalizer was loaded into the neighboring well on the same row as each sample tested, and a blank control (10% FBST only) was included on each biochip; **3)** Secondary antibody incubation: each well was subsequently incubated with a mixture of 4 nM iFluor™820-labeled anti-human IgG secondary antibody, 4 nM CF647-labeled anti-human IgM secondary antibody, and 4 nM iFluor™820-labeled streptavidin in a 2% BSA solution for 30 minutes at room temperature. Each well was washed six times with 1X PBST (10X PBST: 10X PBS with 0.5% Tween 20) between steps. The iFluor™820-labeled streptavidin in the detection step binds to BSA-biotin spots in each well on the pGOLD™ biochip, and the signal was used as an “intrawell signal normalizer”. In parallel, row-dependent, antigen-specific “adjacent well” normalization was performed to normalize antigen printing variation or slide-to-slide variation. Anti-RBD IgG/IgM levels were calculated as the ratio of anti-RBD signal of the sample to that of the adjacent well normalizer and multiplied by 100. Anti-NCP IgG levels were calculated as the ratio of anti-NCP signal of the sample to that of the intrawell normalizer (BSA-Biotin) multiplied by 100.

### pGOLD™ IgG avidity assay

The pGOLD™ IgG avidity index was measured by detecting anti-RBD IgG signals in a dissolved DBS sample in 10% FBST, with and without 4 M urea, respectively. The two solutions were run side-by-side in neighboring pGOLD™ wells, under the otherwise identical pGOLD™ protocol. An avidity index was calculated as the ratio of IgG signal detected in the solution with 4 M urea to that without urea^53^. Avidity is always below the value of 1, since the denaturing urea tends to weaken antibody binding to antigens immobilized on pGOLD™. A higher avidity index corresponds to higher antibody-antigen binding strength.

### Definitions

ROC (receiver operating characteristics) curve analysis was performed based on the quantification of antibody levels of pre-pandemic healthy subjects, vaccinees, and COVID-19 patients by MedCalc 20 (MedCalc Software Ltd, Ostend, Belgium). Cutoff values were determined under a criterion of 100% specificity while maximizing the sensitivity for detecting positive anti-NCP IgG level (> 10.3) in COVID-19 patients and for detecting positive anti-RBD IgG level (> 2.16) in both COVID-19 patients and vaccinated cohorts.

Vaccinated subjects each provided a self-assessment of the severity of side effects after the first and second vaccine doses from a range of 0 (low) to 10 (high). MidaSpot™ COVID-19 antibody rapid test results were visually qualified and defined according to IgM and IgG line intensity as positive or negative (Figure 1).

### Statistical analysis

Continuous values were shown as mean ± standard deviation for normally distributed data and median and interquartile range (IQR) for nonparametric data. The Shapiro □Wilk test was used to determine the normality of anti-RBD IgG levels in each group (COVID-19, pre-pandemic, pre-vaccination D0, D19-28 prior to the second dose, D33-55, D61-83, D89-108, and D112-146). The Mann-Whitney U test was performed to compare anti-RBD IgG levels between two groups for non-parametrically distributed data. A *p* value < 0.05 was considered statistically different between two groups (COVID-19, pre-vaccination D0, D19-28, D33-55, D61-83, D89-108, and D112-146) in this study (OriginPro 2021b, OriginLab Corporation, Northampton, Massachusetts, USA).

## Acknowledgements

This work was supported by Nirmidas Biotech, Inc. internal R&D funds, and by Platelet and Thrombosis Research, LLC, Baltimore, MD, USA.

## Contributions

P.A.G, M.T., and H.D. were involved in study design, interpretation of data, manuscript writing, critical comments, and final approval of the manuscript. T.L., D.S., and J.K. performed the pGOLD™ assay experiments, conducted data analysis, prepared Figures, Tables, and manuscript drafts. J.H., S.Z., and D.Y.S. collected, prepared samples, and performed data analysis. U.S.T and K.P.B were involved in study design, laboratory processing, data collection, statistical analysis, and manuscript preparation. A.R., N.R., and N.W. were involved in patient recruitment and data collection. R.C. and R.E. were involved in study design and manuscript preparation. All authors commented on the manuscript.

## Disclosures

Dr. Gurbel reports grants and personal fees from Bayer HealthCare LLC, Otitopic Inc, Amgen, Janssen, and US WorldMeds LLC; grants from Instrumentation Laboratory, Haemonetics, Medicure Inc, Idorsia Pharmaceuticals, and Hikari Dx; personal fees from UpToDate; Dr Gurbel is a relator and expert witness in litigation involving clopidogrel; in addition, Dr. Gurbel has two patents, Detection of restenosis risk in patients issued and Assessment of cardiac health and thrombotic risk in a patient. Dr. Tantry reports receiving honoraria from UptoDate and Aggredyne. H. Dai worked on this project as a consultant for Nirmidas Biotech Inc., independent of his Stanford projects. Other authors report no disclosures.

## Additional information

Supplementary information is available for this paper.

Correspondence and requests for materials should be addressed to P.A.G, M.T., and H.D.

## SUPPLEMENTARY INFORMATION

**Table S1.**
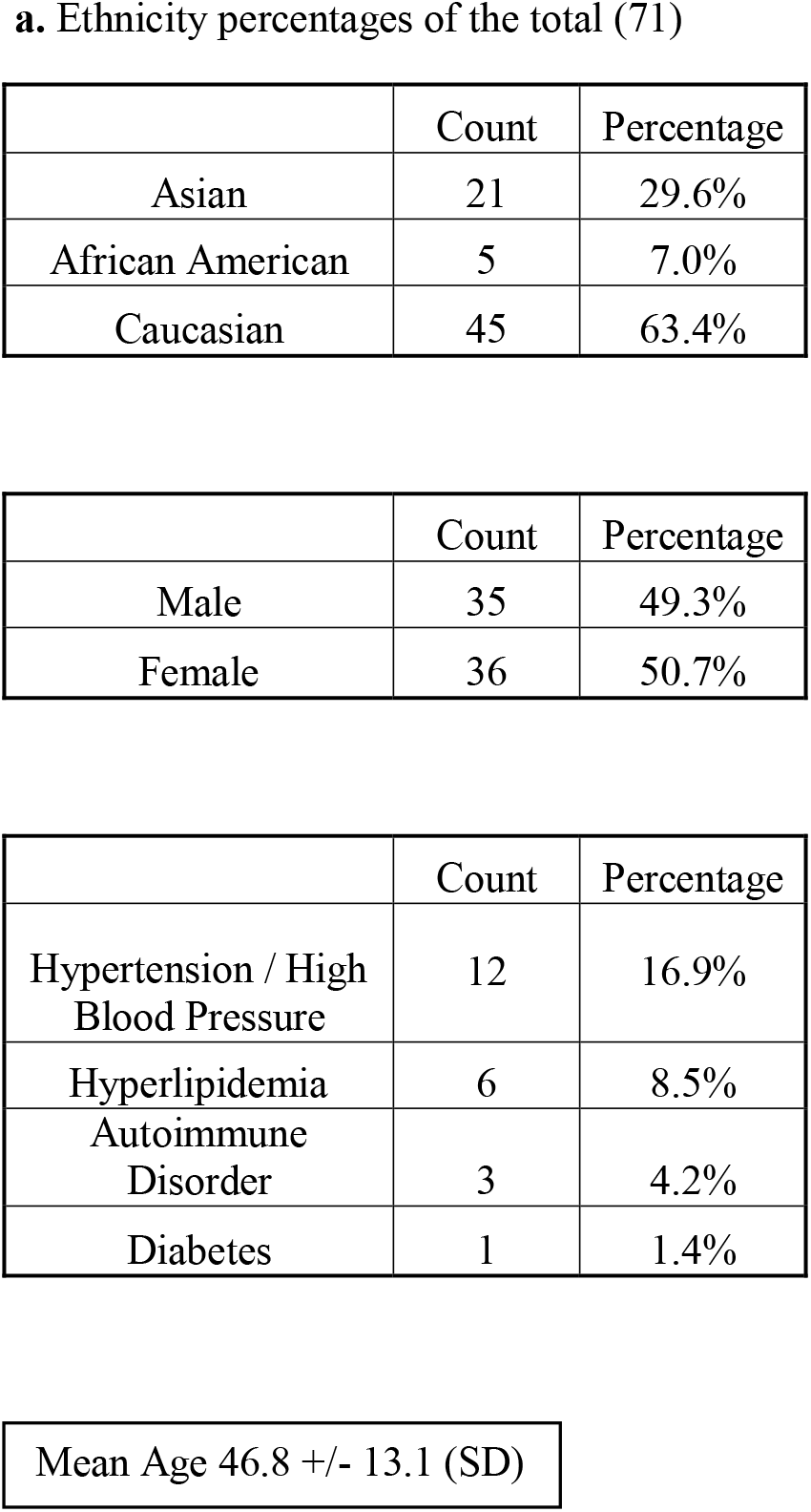
Study subjects’ demographics. (**a**) Caucasians, Asians, and African Americans comprise the 71 subjects of this study. (**b**) An equal gender participation ratio (M/F) is 49.3% vs. 50.7%. (**c**) Hypertension and high blood pressure (HTN/HBP) are the most frequently reported comorbidity of the study participants, at 16.9%. (**d**) The mean age of the study participants is 46.8 years, with a standard deviation of 13.1 years.

**a. Ethnicity percentages of the total (71)**
|  | Count | Percentage |
| --- | --- | --- |
| Asian | 21 | 29.6% |
| African American | 5 | 7.0% |
| Caucasian | 45 | 63.4% |
| Male | 35 | 49.3% |
| Female | 36 | 50.7% |
| Hypertension / High Blood Pressure | 12 | 16.9% |
| Hyperlipidemia | 6 | 8.5% |
| Autoimmune Disorder | 3 | 4.2% |
| Diabetes | 1 | 1.4% |

**Table S1. Study subjects' demographics.**
|  |
| --- |
| Mean Age 46.8 +/- 13.1 (SD) |

**Table S2.** Data summary of anti-RBD IgG levels in each group. The SARS-CoV-2 infected group refers to individuals who provided DBS samples for analysis of anti-RBD and anti-NCP IgG levels after symptom onset and after confirmatory tests were conducted. D0 is the group of individuals who provided DBS samples one day prior to their first dose of vaccine. D19-28, D33-55, D61-83, D89-108, and D112-146 refer to the timepoints at which the vaccinated study participants provided samples for analysis. The prepandemic data are from serum samples collected prior to November 2019. Minimum, Q1, median, Q3, and maximum IgG Levels refer to the pGOLD™ anti-RBD results obtained after performing analysis of the samples from each of these groups. Testing was performed at Nirmidas Biotech, Inc. in Palo Alto, California. Statistical analysis was performed using OriginPro 2021b.

| Grouping on Vaccinated Time or Infection Status | Group Size | Minimum Anti-RBD IgG Level | 25<sup>th</sup> Percentile Anti-RBD IgG Level | Median Anti-RBD IgG Level | 75<sup>th</sup> Percentile Anti-RBD IgG Level | Maximum Anti-RBD IgG Level |
| --- | --- | --- | --- | --- | --- | --- |
| SARS-CoV-2 Infected | 34 | 0.95 | 18.785 | 44.515 | 123.335 | 263.39 |
| D0 pre-vaccination | 23 | -0.05 | 0.01 | 0.07 | 0.93 | 2.18 |
| D19-28 post-first dose | 32 | 2.29 | 11.51 | 21.45 | 33.4825 | 257.82 |
| D33-55 post-first dose | 35 | 50.1 | 103.31 | 188 | 271 | 811 |
| D61-83 post-first dose | 50 | 16.62 | 47.82 | 105.94 | 150.2625 | 308.94 |
| D89-108 post-first dose | 15 | 16.52 | 59.44 | 79.62 | 162.26 | 185.53 |
| D112-146 post-first dose | 43 | 11.3 | 27.68 | 55.98 | 90 | 213.44 |
| Pre-pandemic | 200 | -0.11 | -0.02 | 0 | 0.1725 | 2.16 |

**Table S3.** Data normality (Shapiro-Wilk) test of Anti-RBD IgG levels for each group. This table describes the sample size and normality status defining characteristics of 200 pre-pandemic serum samples, and the DBS samples collected at different timepoints pre- and post-vaccination of the 71 study participants. D89-108 data is normally distributed, with all the other groups having non-normally distributed IgG levels. Statistical analysis was performed using OriginPro 2021b.

| Study Group | Group Size | *p* value | Statistic | Decision at level (5%) |
| --- | --- | --- | --- | --- |
| Pre-pandemic | 200 | 0 | 0.59435 | Reject normality |
| Day 0 | 23 | 0.0002 | 0.79319 | Reject normality |
| Day 19-28 | 32 | <0.0001 | 0.51508 | Reject normality |
| Day 33-55 | 35 | <0.0001 | 0.81345 | Reject normality |
| Day 61-83 | 50 | 0.00239 | 0.92014 | Reject normality |
| Day 89-108 | 15 | 0.27743 | 0.93048 | Cannot reject normality |
| Day 112-146 | 43 | 0.0005 | 0.88812 | Reject normality |

**Table S4.**
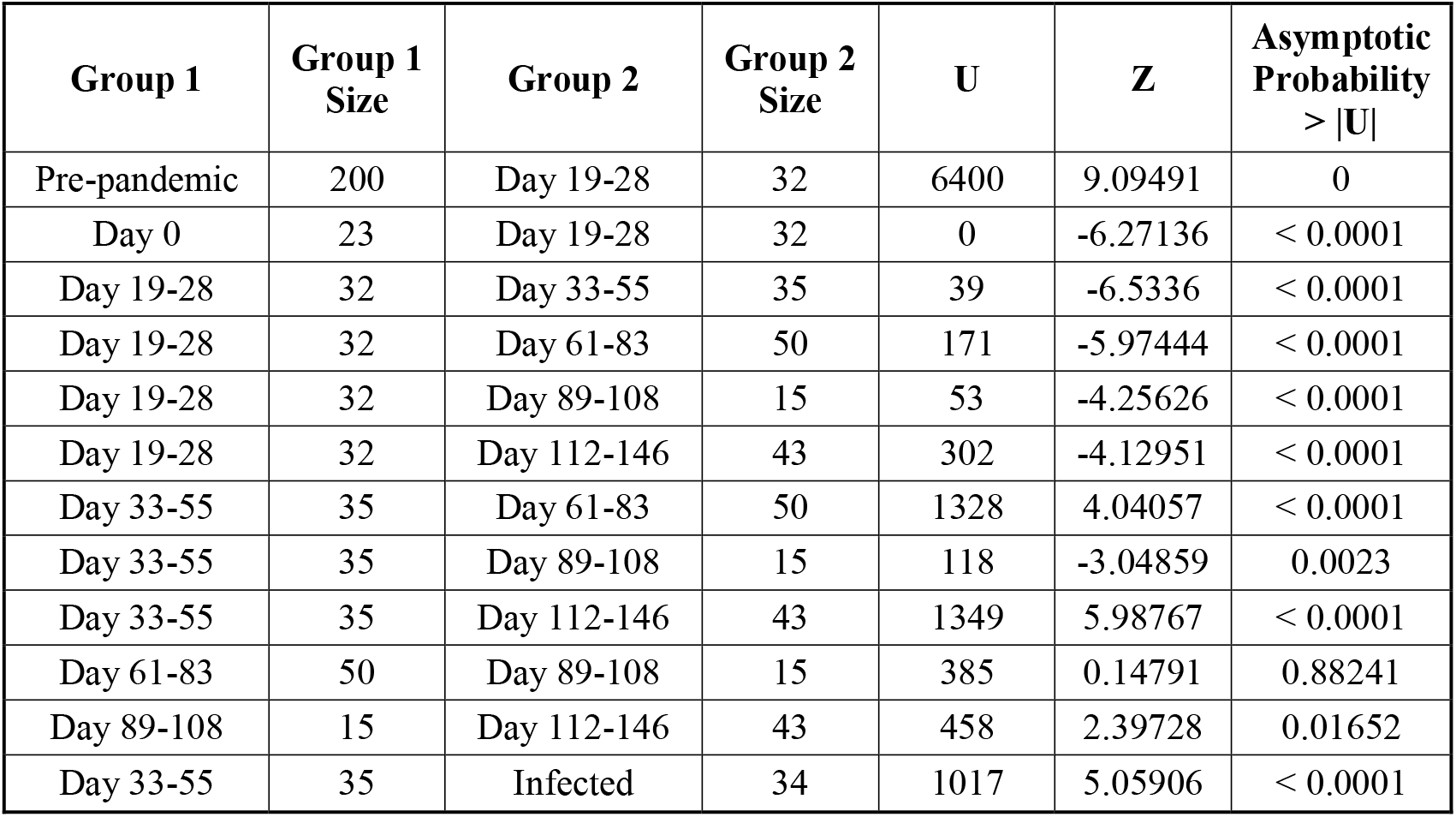
Comparison of Anti-RBD IgG levels between groups by the Mann-Whitney Test. Comparing anti-RBD IgG levels between different groups by the Mann-Whitney test indicated the similarity between groups, according to the asymptotic probability values. *p* > 0.05 indicates similarity between two groups; Otherwise, *p* < 0.05 indicates a clear difference between two groups with statistical significance. Statistical analysis was performed using OriginPro 2021b.

| Group 1 | Group 1 Size | Group 2 | Group 2 Size | U | Z | Asymptotic Probability > \|U\| |
| --- | --- | --- | --- | --- | --- | --- |
| Pre-pandemic | 200 | Day 19-28 | 32 | 6400 | 9.09491 | 0 |
| Day 0 | 23 | Day 19-28 | 32 | 0 | -6.27136 | < 0.0001 |
| Day 19-28 | 32 | Day 33-55 | 35 | 39 | -6.5336 | < 0.0001 |
| Day 19-28 | 32 | Day 61-83 | 50 | 171 | -5.97444 | < 0.0001 |
| Day 19-28 | 32 | Day 89-108 | 15 | 53 | -4.25626 | < 0.0001 |
| Day 19-28 | 32 | Day 112-146 | 43 | 302 | -4.12951 | < 0.0001 |
| Day 33-55 | 35 | Day 61-83 | 50 | 1328 | 4.04057 | < 0.0001 |
| Day 33-55 | 35 | Day 89-108 | 15 | 118 | -3.04859 | 0.0023 |
| Day 33-55 | 35 | Day 112-146 | 43 | 1349 | 5.98767 | < 0.0001 |
| Day 61-83 | 50 | Day 89-108 | 15 | 385 | 0.14791 | 0.88241 |
| Day 89-108 | 15 | Day 112-146 | 43 | 458 | 2.39728 | 0.01652 |
| Day 33-55 | 35 | Infected | 34 | 1017 | 5.05906 | < 0.0001 |

**Table S5.** Samples with negative MidaSpot™ results compared to pGOLD™ values. This table presents the samples which were expected to have positive MidaSpot™ results but were negative. This data compares the MidaSpot™ results to the pGOLD™ values, which show that the pGOLD™ anti-RBD IgG levels of the same samples are near the minimum values in their groups. Sample AbV-32 at the previous timepoint had a low IgG signal on MidaSpot™ and further became faintly positive on MidaSpot™ at the D112-146 timepoint.

| Samples negative on MidaSpot™ corresponding to their pGOLD™ results |  |  |  |  |  |
| --- | --- | --- | --- | --- | --- |
| Sample ID | Day Post First Dose Vaccine (Group) | MidaSpot™ IgG result | pGOLD™ anti-RBD IgG Level for the Sample | pGOLD™ Minimum anti-RBD IgG Level in Group | pGOLD™ Median anti-RBD IgG Level in Group |
| AbV-22 | 121 (D112-146) | neg | 14.25 | 11.30 | 55.98 |
| AbV-32 | 67 (D63-81) | neg | 18.97 | 16.62 | 105.94 |
| AbV-36 | 118 (D112-146) | neg | 18.81 | 11.30 | 55.98 |

## Notes

### Clinical Trial

NCT04910971

### Funding Statement

This work was supported by Nirmidas Biotech. Inc. internal R&D funds and funds from Platelet and Thrombosis Research, LLC, Baltimore, MD, USA

### Author Declarations

LifeBridge Health local institutional review board under Protocol # 1707882

